# Risk Prediction for Non-cardiac Surgery Using the 12-Lead Electrocardiogram: An Explainable Deep Learning Approach

**DOI:** 10.1101/2024.11.19.24317577

**Authors:** Carl Harris, Anway Pimpalkar, Ataes Aggarwal, Jiyuan Yang, Xiaojian Chen, Samuel Schmidgall, Sampath Rapuri, Joseph L. Greenstein, Casey Overby Taylor, Robert D. Stevens

## Abstract

**Background:** To improve on existing noncardiac surgery risk scores, we propose a novel approach which leverages features of the preoperative 12-lead electrocardiogram (ECG) to predict major adverse postoperative outcomes.

**Methods:** Data acquired in 37,060 adult patients prior to major noncardiac surgery were used to train a series of convolutional neural network models in the task of predicting in-hospital acute myocardial infarction (MI), in-hospital mortality (IHM), and a composite of in-hospital MI, in-hospital stroke, and 30-day mortality. Preoperative ECG waveform features were first modeled as sole inputs then integrated with clinical variables in fusion models. Model discrimination was evaluated using area under the receiver operating characteristic (AUROC) analysis, and performances were compared with the Revised Cardiac Risk Index (RCRI), a benchmark preoperative risk score To gain interpretable insight, a generative approach using counterfactual ECGs was implemented.

**Results:** The ECG fusion model had an AUROC of 0.858 (95% CI [0.845, 0.872]), 0.899 (95% CI [0.889, 0.908]), and 0.835 (95% CI [0.827, 0.843]) in predicting MI, IHM, and the composite endpoint, respectively; these AUROC values were significantly higher than in models based on ECG waveforms alone (MI: *p* = 0.001, IHM: *p* < 10^−4^, composite: *p* < 10^−4^). All ECG based models had significantly higher discrimination than the RCRI. Counterfactual ECG analysis revealed morphological features relevant to outcome classification.

**Conclusion:** A deep learning approach integrating preoperative ECG waveform features significantly enhances the ability to predict major outcomes after noncardiac surgery. The use of counterfactual ECGs provides plausible explanations for classifier decisions, increasing the interpretability of the models.

**Clinical perspective:** What is new?

□ A deep learning approach applied to preoperative 12-lead ECG waveforms accurately predicts major outcomes after noncardiac surgery.
□ This model outperforms the benchmark Revised Cardiac Risk Index (RCRI).
□ The highest predictive performance was obtained with a fusion model that combines preoperative ECG waveforms with routinely collected clinical variables.
□ An exploratory approach in which counterfactual ECGs are generated provides explainability for classifier decisions.

What are the clinical implications?

□ In adults undergoing non-cardiac surgery, ECG waveform features are predictive of postoperative cardiovascular risk
□ Risk models integrating ECG waveforms with clinical variables can serve as the basis for outcome modifying interventions across the surgical continuum.
□ Achieving explainability through counterfactual ECGs represents an important step towards real-world implementation.

## Introduction

Despite advances in perioperative safety, adult patients undergoing surgery continue to incur Major Adverse Cardiovascular and Cerebrovascular Events (MACCE) such as myocardial infarction (MI) and ischemic stroke, with an incidence of MACCE after noncardiac surgery reported between 1 and 7 percent depending on the population studied^1–3^. These complications prolong hospitalization^4^, increase medical costs^5^, and may burden surviving patients with disabilities that reduce their quality of life in the long term^6^. Accurate preoperative risk stratification^7^ may have actionable contingencies^8^. Medical conditions such as coronary artery disease, dysrhythmias, hypertension, or diabetes mellitus can be optimized prior to surgery^9^, while the invasiveness of surgery, type of anesthesia^10^ and the intensity and duration of perioperative monitoring can be customized ^11^; moreover, appropriately risk stratified patients can make informed decisions prior to agreeing to an operation^12^. However, existing preoperative risk stratification tools have only modest discrimination. The Revised Cardiac Risk Index (RCRI), a score regarded as a benchmark in predicting cardiac events after noncardiac surgery, has a median area under the receiver-operating curve (AUROC) of 0.75, sensitivity of 0.65, and specificity of 0.76 according to a meta-analysis of 18 studies^13^. There is consequently an unmet need for more accurate tools for risk stratification.

The 12-lead ECG is widely used to diagnose a range of cardiac conditions such as dysrhythmias, conduction abnormalities, or evidence of ischemic heart disease^14,15^. Recent studies indicate that the ECG may also be valuable in predicting future events such as paroxysmal atrial fibrillation^16–18^ or sudden cardiac death^19,20^. This research leverages very large datasets of 12-lead ECGs to train supervised machine learning algorithms. The central conjecture in these studies is that ECG waveforms contain previously unidentified predictive features which might be characterized as latent because not recognizable even by trained clinical practitioners. In this work, we reasoned that a similar paradigm might be relevant in the prediction of postoperative outcomes. Using preoperative 12-lead ECGs, we trained a deep learning model to predict the risk of in-hospital MI, in-hospital mortality, and a composite endpoint of in-hospital MI, in-hospital stroke, and 30-day mortality among patients undergoing major noncardiac surgery.

## Methods

### Objectives

The primary objective is to predict major adverse postoperative events using preoperative 10- second, 12-lead ECG (**Figure 1A**). We identified three postoperative endpoints of interest: myocardial infarction (MI), in-hospital mortality (IHM), and a composite of stroke, MI, and mortality (composite). MI and stroke were identified using International Classification of Diseases (ICD) 9 and 10 codes (see **Table S1**). IHM was derived from a reference flag for whether the patient died in the hospital, and 30-day mortality was extrapolated from a combination of in-hospital records and state death records. We investigated our task through two modeling approaches. First, we developed a deep learning model to predict adverse postoperative outcomes in noncardiac surgical patients using ECG waveforms alone (WF model). And second, we devised a fusion model that combines ECG waveform data with routinely collected clinical variables to predict these outcomes (Fusion model). Our first analysis examines whether there are latent factors within the ECG that are predictive of the outcomes of interest; the second allows us to determine if the ECG contains information complementary to structured electronic healthcare record (EHR) data that increases accuracy of prediction of postoperative outcomes as compared to structured data alone.

**Figure 1.**
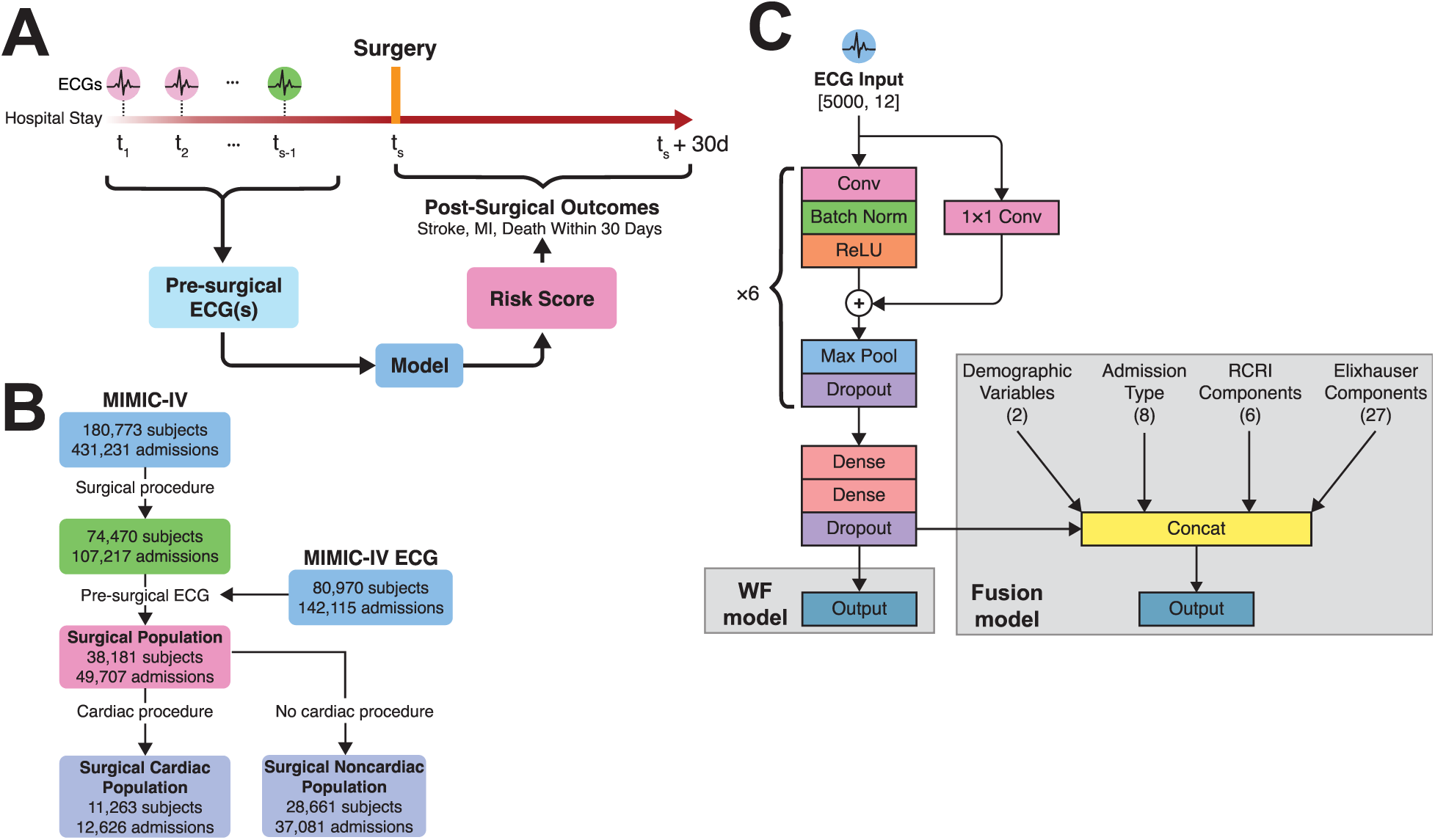
Task of interest, selection criteria, and model architecture. **(A)** Schematic of our task of interest, which is to predict adverse postoperative outcomes using preoperative ECGs. **(B)** Flowchart used for patient selection. **(C)** Model architecture. We leveraged a CNN backbone to make predictions based on preoperative ECGs alone (WF model) or in combination with routinely collected clinical and demographic variables available preoperatively (fusion model).

### Dataset

Data were extracted from MIMIC-IV (v2.2)^21,22^, which includes 299,712 patients across 431,231 admissions to Beth Israel Deaconess Medical Center between 2008 and 2019. We included adult patients undergoing major noncardiac surgery identified via procedure ICD codes recorded during their stay, using Procedure Classes groupings provided by the Healthcare Cost and Utilization Project^23^. This designates all ICD-9 and ICD-10 procedure codes into four categories, based on whether the procedure is minor (non-operating room) or major (operating room) and whether it is diagnostic or therapeutic. Surgical patients were identified by those who underwent any major procedure according to the ICD codes provided. Next, we differentiated between major cardiac and noncardiac surgeries. Again, we used the Clinical Classification Software (CCS) – patients who first underwent a major cardiac surgery were considered cardiac patients; patients whose first procedures were noncardiac were classified as noncardiac surgical patients (**Figure 1B**). A summary of patient characteristics can be found in **Table 1**.

**Table 1.**
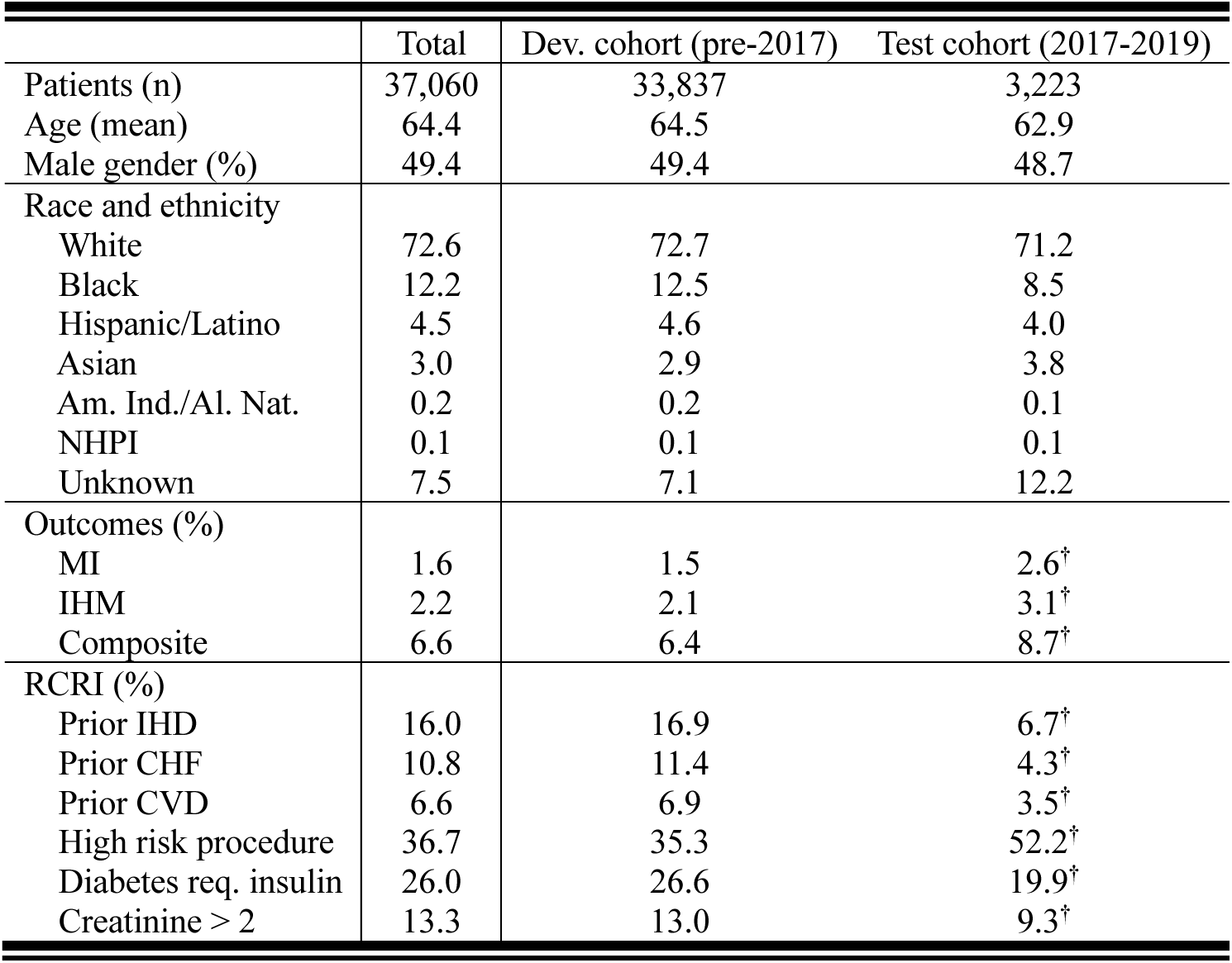
Population sample characteristics. Shown are demographic, outcome, and RCRI components for the overall sample, and our temporally stratified one (see **Methods**). The † symbol denotes significant differences (*p* < 10^−3^) between the development cohort and test cohort, based on a normal test for proportion differences in unpaired samples^24^.

|  | Total | Dev. cohort (pre-2017) | Test cohort (2017-2019) |
| --- | --- | --- | --- |
| Patients (n) | 37,060 | 33,837 | 3,223 |
| Age (mean) | 64.4 | 64.5 | 62.9 |
| Male gender (%) | 49.4 | 49.4 | 48.7 |
| Race and ethnicity |  |  |  |
| White | 72.6 | 72.7 | 71.2 |
| Black | 12.2 | 12.5 | 8.5 |
| Hispanic/Latino | 4.5 | 4.6 | 4.0 |
| Asian | 3.0 | 2.9 | 3.8 |
| Am. Ind./Al. Nat. | 0.2 | 0.2 | 0.1 |
| NHPI | 0.1 | 0.1 | 0.1 |
| Unknown | 7.5 | 7.1 | 12.2 |
| Outcomes (%) |  |  |  |
| MI | 1.6 | 1.5 | 2.6<sup>†</sup> |
| IHM | 2.2 | 2.1 | 3.1<sup>†</sup> |
| Composite | 6.6 | 6.4 | 8.7<sup>†</sup> |
| RCRI (%) |  |  |  |
| Prior IHD | 16.0 | 16.9 | 6.7<sup>†</sup> |
| Prior CHF | 10.8 | 11.4 | 4.3<sup>†</sup> |
| Prior CVD | 6.6 | 6.9 | 3.5<sup>†</sup> |
| High risk procedure | 36.7 | 35.3 | 52.2<sup>†</sup> |
| Diabetes req. insulin | 26.0 | 26.6 | 19.9<sup>†</sup> |
| Creatinine > 2 | 13.3 | 13.0 | 9.3<sup>†</sup> |

Within MIMIC-IV, the timing of procedures was identified at the granularity of a day – this means that for ECGs recorded on the same day as a procedure, we could not conclusively determine if the ECG was administered before the procedure or after it. Because our task of interest involves predicting outcomes using preoperative ECGs, we only included ECGs obtained at least one day before surgery.

### Evaluation

We adopted two evaluation approaches. In the first, we used a standard *k*-fold validation with 10 folds. We randomly divided the dataset into 10 partitions, split by subject (so that no subject could be in both the train and test partitions). Then, for each partition, we trained on the nine other partitions and evaluated on the held-out test partition. Statistical analysis was then calculated across the 10 test folds. In the second evaluation method, we trained and tested our method on chronologically distinct cohorts. Specifically, we trained our model on patients admitted between 2008 and 2016 and tested our model on a cohort admitted between 2017 and 2019. The goal of the second evaluation strategy was to simulate a single-site sequential validation, as motivated by Sundrani et al.^25^.

### Models

We developed two models to predict outcomes (**Figure 1C**). The WF model used only preoperative ECGs to predict postoperative outcomes. We selected a 1D convolutional neural network (CNN) as our model backbone, based in part on the architecture proposed by Attia et al.^26^. This architecture takes in a raw 12-lead, 10-second ECG sampled at 500 Hz (i.e., a [5000, 12]-dimensional array of voltages) and is trained to output an estimated probability of a given condition *ŷ* ∈ [0,1]. The core of the network is six blocks of convolutional layers. Each block consists of a 1D convolutional layer, followed by a batch normalization layer and an activation layer with rectified linear unit (ReLU) activation. We also employed residual connections in each convolutional block, which allows for gradients to better pass through to earlier layers of the network^27^. The residual layer uses a 1 × 1 convolution to match the dimensionality of the output of the main convolutional layer. We used filter numbers [16, 16, 32, 32, 64, 64] and corresponding kernel widths [7, 7, 5, 5, 3, 3] in the convolutional layers across the six convolutional blocks. Following the convolutional blocks are two dense layers (hidden sizes of 64 and 32, respectively), and then a dropout layer. After the second dense layer we used an output layer with sigmoid activation, which predicts a risk of the outcome (e.g., MI), *ŷ*, where *ŷ* ∈ [0,1]. An explanation of our hyperparameter optimization process can be found in the **Supplementary Methods** and **Table S2**.

The second model we introduced is a fusion model, which integrates ECG waveform information with clinical risk factors. Specifically, we combined the CNN backbone with basic demographic information (age and sex), admission type (e.g., transfer, emergency, elective, etc.), binary indicators for each of the six components of the RCRI, and 26 binary indicators from the Elixhauser Comorbidity Score (ECS). ECS components were identified via ICD-9 and ICD-10 codes^28^. Because ICD codes are only tabulated at the *end* of patients’ stays, it was imperative we did not include as input features conditions that could reasonably have resulted from the surgery. For this reason, we excluded congestive heart failure, cardiac arrythmias, coagulopathy, and blood loss anemia from the set of components we include in our fusion model. We also excluded HIV status as these data are not available in MIMIC-IV. A full list of included variables is described in the **Supplementary Methods**.

### Model training

We implemented the waveform and fusion models in TensorFlow (version 2.15.0) and the counterfactual model in PyTorch (version 2.1.1). We trained models for up to 100 epochs using an Adam optimizer with an initial learning rate of 1 × 10^−3^. We randomly selected 10% of patients (and their corresponding ECGs) for validation and calculated the validation loss at each epoch. If the validation loss did not decrease for three consecutive epochs, we reduced the learning rate by a factor of 0.5. If the validation loss did not decrease for six epochs, training terminated, and the weights corresponding to the minimum validation loss were restored. We applied minimal preprocessing to our data. ECGs were dropped if any entries within the ECG waveform were invalid (< 2% of all ECGs). In the fusion model, we applied standard scaling to age but left the rest of the binary variables unscaled.

### Model performance

Our primary metric of interest was area under the receiver-operating curve (AUROC). In addition, we also calculated the threshold-independent area under the precision-recall curve (AUPRC). For threshold-dependent measures (sensitivity, specificity, PPV, NPV, OR) we set a risk score cutoff for high and low risk as *ŷ* = 0.05; for RCRI, we followed previous literature^29,30^ and denoted high risk patients as those with a composite RCRI score ≥ 2. For each model, we calculated the risk score for all preoperative ECGs (e.g., if a patient had five preoperative ECGs, each was fed into the model at inference time to produce a risk score *ŷ*). We then selected the maximum risk score across all preoperative ECGs for a given patient as that patient’s risk of an adverse postoperative event.

In comparing our approach to the RCRI, we also computed the Net Reclassification Index^31^ (NRI) between our models and RCRI. Briefly, the NRI measures how well a new model (e.g., WF and fusion models) reclassifies subjects relative to a baseline (RCRI). The NRI quantifies the correct upward or downward movement in risk categories for individuals, and its value can range from −2 to 2. Positive values indicate an improvement in classification with the new model, while negative values suggest a deterioration (see the **Supplementary Methods**).

### Statistical analysis

To generate confidence intervals, we relied on a bootstrapping approach with 10,000 iterations. For statistical tests, we applied a permutation test with 10,000 iterations, followed by a Bonferroni correction^32^ to establish statistical significance given multiple comparisons.

### Model comparisons

Our primary comparison benchmark was the RCRI, which consists of six components: preoperative creatinine > 2 mg/dL, a diagnosis of diabetes mellitus requiring insulin, whether the surgery was high risk (intraperitoneal, intrathoracic, suprainguinal vascular), and whether the patient had a history of ischemic heart disease, congestive heart failure, or cerebrovascular disease (see **Supplementary Methods**). Each of these conditions was coded as a binary variable, and their sum was the patient’s RCRI score (in the range from 0 to 6).

In addition, we compared our WF and fusion models to one trained using ECI variables, as well as three CNN-based models from Hannun et al.^33^, Ribiero et al.^34^, and Ouyang et al.^35^, trained using the same training and evaluation schemes and data as our model. In addition, we implemented a non-DL based ECG classifier based on the machine-generated features from the ECG hardware (see **Supplementary Methods**).

### Explainability

To identify the characteristics relevant to classification decisions, we proposed a generative approach which creates counterfactual ECGs. Briefly, following the training paradigm previous described, we have a classifier that, given an input ECG, produces a risk score (e.g., the probability of postoperative MI). Our counterfactual model seeks to intervene on this input waveform by introducing some minimal, physiologically plausible modifications to the underlying morphology such that it elicits a different, user-defined risk score *δ* ∈ [0, 1] (see **Supplementary Methods**). For example, given an ECG with a low classifier-assigned risk score (e.g., a 0.1% risk of MI), we could generate counterfactual versions of this ECG such that it is classified at medium (e.g., *δ* = 0.03, 3% risk of MI) or high (e.g., *δ* = 0.15, 15% risk of MI) risk by modifying the classification-decision relevant areas of the ECG waveform such that the classifier judges the counterfactual to be at the prescribed risk level. A successful counterfactual ECG demonstrates three characteristics^36,37^: (1) it will appear similar to the input (i.e., be minimally modified relative to the original ECG); (2) it will elicit the desired risk score *δ* from the classifier; and (3) it will remain in-distribution (i.e., “look like” a real ECG). To enforce these criteria, we used reconstruction losses (i.e., *ℓ*_2_) to ensure the simulated ECG was similar to the input (satisfying 1), a KL-divergence based loss to encourage the generator to create ECGs that elicit the desired risk score *δ* from the classifier (satisfying 2), and a generative adversarial network (GAN) to ensure the synthetic ECGs remain in-distribution (satisfying 3). Given these criteria, the counterfactual model could then be leveraged for explainability – we could create counterfactual ECGs at varying risk levels, allowing for the identification of features driving classifier decisions. By comparing high- and low-risk counterfactuals, we would be able to capture, visually and empirically, morphological characteristics which the algorithm considers in accomplishing the classification task.

To substantiate our claim that the counterfactual approach identified relevant predictive characteristics, we used a separate diagnostic dataset. This dataset consists of 12-lead ECGs from 45,152 patients derived from Chapman University, Shaoxing People’s Hospital, and Ningbo First Hospital and was labeled by medical experts^38,39^. The goal of this analysis was to train counterfactual models to predict obvious and well-defined conditions (e.g., atrial flutter), and demonstrate that our model replicates these conditions through visual inspection and empirical investigation. We did this first in a diagnostic dataset, where morphological characteristics are well-established, and then applied it to the prognostic dataset as a feature exploration exercise (where ground-truth characteristics for “high risk of future MI,” for example, are unknown or poorly defined).

## Results

The analysis was conducted on 37,060 adult patients undergoing noncardiac surgery, with a mean age of 64.4 years, of whom 49.4% were male. Comorbid conditions were common, with notable prevalences of prior ischemic heart disease (16.0%), diabetes requiring insulin (26.0%), and elevated creatinine (13.3%). Approximately 36.7% of procedures were high-risk (intraperitoneal, intrathoracic, and suprainguinal vascular surgery^29^). Racial and ethnic demographics show the majority of patients are White (72.6%), with representation from Black (12.2%) and Hispanic/Latino (4.5%) groups.

### Model performance

Model performance for the cross validation and temporal stratification is shown in **Tables 2** and **3**, respectively. We compare our WF and fusion models to six baselines – RCRI, a model trained solely on the Elixhauser components, three other CNN architectures (PreOpNet, Hannun et al., and Ribeiro et al.), as well as a non-deep learning approach based on simple, machine-extracted ECG features (e.g., RR-interval, QRS axis, etc.). Our primary performance metric of interest is AUROC; unless otherwise noted, we follow the format: AUROC (95% CI [lower, upper]), where confidence intervals are determined via our bootstrapping approach. Performance from cross-validation and temporal stratification is shown in **Figure 2**.

**Figure 2.**
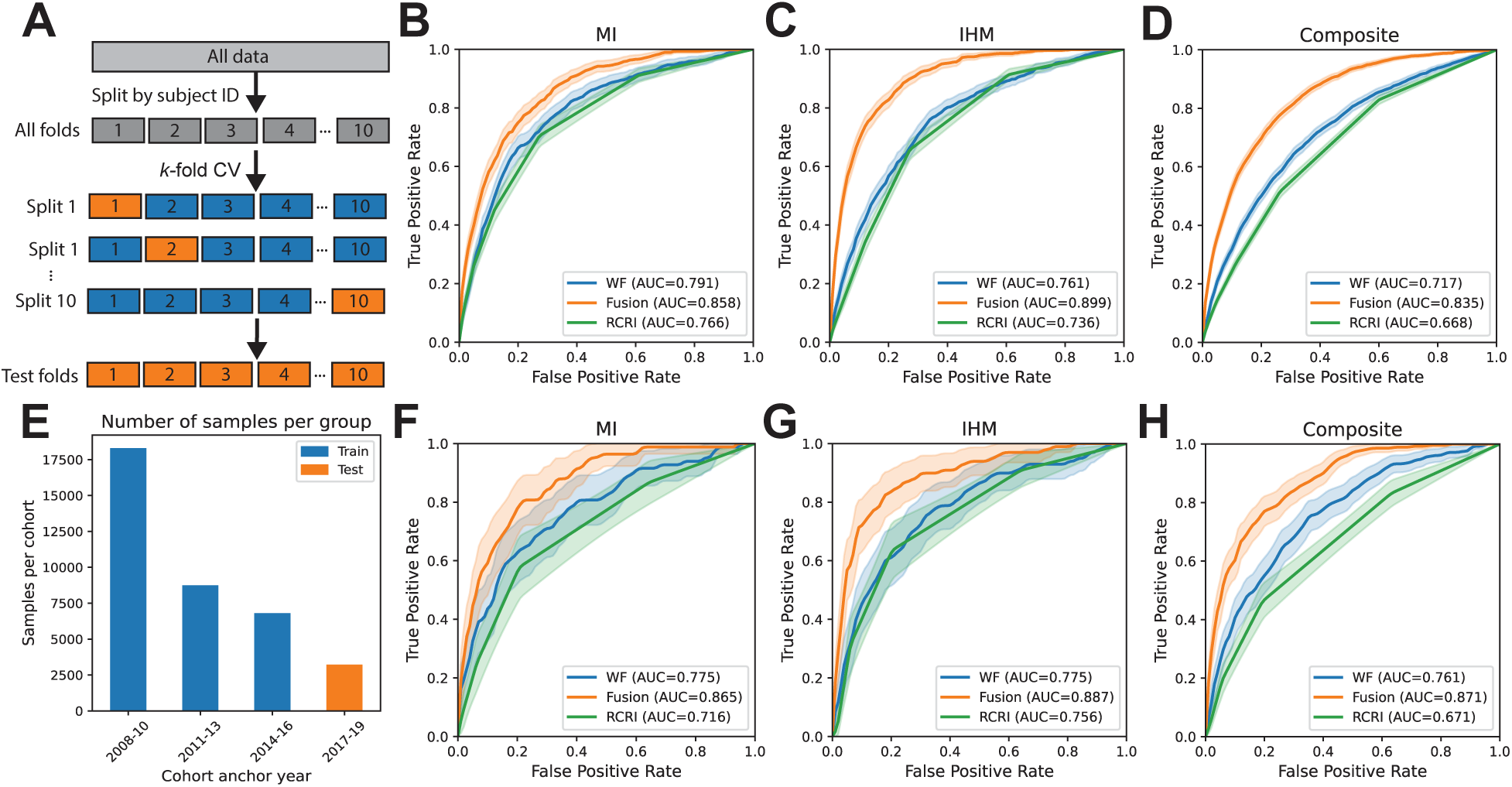
Model performance. **(A)** Schematic of the k-fold validation scheme for the main results, where data are split into 10 folds (by patient ID). Train folds are indicated in blue, and test folds in orange. Test folds are combined to evaluate the final performance of the model, as shown by the ROC curves (with AU-ROC values inset) in predicting postoperative **(B)** MI, **(C)** IHM, and **(D)** composite outcome. Shaded areas denote 95% confidence intervals from 10,000 bootstrapped samples. **(E)** Number of samples in the temporal stratification analysis. Blue bars are used to train the model (corresponding to patients admitted between 2008 and 2016), and samples corresponding to the orange bar (2017-19) are used to evaluate it. The corresponding performance is shown in panels **(F)**, **(G)**, and **(H)**. Corresponding AUPRC plots are shown in **Figure S1**.

**Table 2.** Cross-validation model comparison. Table shows the performance of different models across the three outcomes of interest. We compare the performance of our model (both waveform only and fusion) with our SOC baseline (RCRI), a model trained using only Elixhauser components, three other CNN architectures (PreOpNet^35^, Hannun et al.^33^, and Ribeiro et al.^34^) as well as a traditional ML model based on ECG features and an XGBoost classifier^41^.“AUROC” and “AUPRC” denote the area under the receiver operator and precision recall curves, respectively. “PPV” and “NPV” denote the positive and negative predictive value; “OR” denotes the odds-ratio. Threshold-dependent measures (“sensitivity”… “OR”) are calculated using a threshold of 0.05 for the ML algorithms, and a threshold of 2 for RCRI.

| MI |  |  |  |  |  |  |  |
| --- | --- | --- | --- | --- | --- | --- | --- |
| Model | AUROC | AUPRC | Sensitivity | Specificity | PPV | NPV | OR |
| WF Model | 0.791<br>[0.771, 0.809] | 0.067<br>[0.056, 0.082] | 0.558<br>[0.518, 0.599] | 0.857<br>[0.854, 0.861] | 0.059<br>[0.053, 0.065] | 0.992<br>[0.991, 0.993] | 7.21<br>[6.14, 8.51] |
| Fusion model | 0.858<br>[0.845, 0.872] | 0.122<br>[0.103, 0.146] | 0.580<br>[0.540, 0.620] | 0.900<br>[0.897, 0.903] | 0.085<br>[0.076, 0.093] | 0.993<br>[0.992, 0.993] | 11.45<br>[9.78, 13.44] |
| RCRI | 0.766<br>[0.747, 0.784] | 0.051<br>[0.044, 0.060] | 0.705<br>[0.668, 0.742] | 0.726<br>[0.722, 0.731] | 0.040<br>[0.036, 0.043] | 0.994<br>[0.993, 0.994] | 6.14<br>[5.15, 7.37] |
| Elixhauser comp. | 0.790<br>[0.771, 0.808] | 0.072<br>[0.061, 0.087] | 0.481<br>[0.440, 0.522] | 0.891<br>[0.888, 0.894] | 0.066<br>[0.059, 0.074] | 0.991<br>[0.990, 0.992] | 7.15<br>[6.08, 8.38] |
| Hannun et al.<sup>33</sup> | 0.771<br>[0.751, 0.791] | 0.060<br>[0.051, 0.073] | 0.502<br>[0.461, 0.542] | 0.869<br>[0.866, 0.873] | 0.058<br>[0.051, 0.064] | 0.991<br>[0.990, 0.992] | 6.36<br>[5.41, 7.46] |
| Ribiero et al.<sup>34</sup> | 0.750<br>[0.730, 0.770] | 0.062<br>[0.052, 0.078] | 0.420<br>[0.380, 0.459] | 0.880<br>[0.876, 0.883] | 0.053<br>[0.047, 0.059] | 0.990<br>[0.988, 0.991] | 5.06<br>[4.31, 5.93] |
| PreOpNet<sup>35</sup> | 0.715<br>[0.694, 0.736] | 0.037<br>[0.033, 0.044] | 0.913<br>[0.889, 0.935] | 0.264<br>[0.260, 0.269] | 0.019<br>[0.018, 0.021] | 0.995<br>[0.993, 0.996] | 3.70<br>[2.86, 5.08] |
| Machine features | 0.687<br>[0.665, 0.710] | 0.038<br>[0.033, 0.046] | 0.327<br>[0.289, 0.366] | 0.887<br>[0.884, 0.890] | 0.044<br>[0.038, 0.050] | 0.988<br>[0.987, 0.989] | 3.69<br>[3.09, 4.36] |
| IHM |  |  |  |  |  |  |  |
| WF Model | 0.761<br>[0.744, 0.777] | 0.071<br>[0.063, 0.082] | 0.582<br>[0.549, 0.615] | 0.788<br>[0.784, 0.792] | 0.059<br>[0.054, 0.064] | 0.988<br>[0.987, 0.989] | 4.92<br>[4.31, 5.64] |
| Fusion model | 0.899<br>[0.889, 0.908] | 0.192<br>[0.171, 0.218] | 0.727<br>[0.695, 0.756] | 0.884<br>[0.881, 0.888] | 0.125<br>[0.116, 0.134] | 0.993<br>[0.992, 0.994] | 17.92<br>[15.46, 20.91] |
| RCRI | 0.736<br>[0.721, 0.752] | 0.050<br>[0.045, 0.057] | 0.657<br>[0.625, 0.689] | 0.728<br>[0.723, 0.733] | 0.052<br>[0.048, 0.056] | 0.989<br>[0.988, 0.991] | 4.91<br>[4.28, 5.68] |
| Elixhauser comp. | 0.834<br>[0.819, 0.850] | 0.136<br>[0.120, 0.156] | 0.611<br>[0.577, 0.644] | 0.885<br>[0.881, 0.888] | 0.107<br>[0.099, 0.116] | 0.990<br>[0.989, 0.991] | 10.87<br>[9.49, 12.46] |
| Hannun et al.<sup>33</sup> | 0.746<br>[0.729, 0.762] | 0.063<br>[0.056, 0.072] | 0.464<br>[0.430, 0.499] | 0.855<br>[0.851, 0.858] | 0.068<br>[0.061, 0.074] | 0.986<br>[0.985, 0.987] | 4.81<br>[4.21, 5.51] |
| Ribiero et al.<sup>34</sup> | 0.723<br>[0.706, 0.740] | 0.058<br>[0.051, 0.067] | 0.428<br>[0.394, 0.462] | 0.839<br>[0.836, 0.843] | 0.057<br>[0.051, 0.063] | 0.985<br>[0.983, 0.986] | 3.73<br>[3.25, 4.27] |
| PreOpNet<sup>35</sup> | 0.659<br>[0.640, 0.678] | 0.045<br>[0.039, 0.052] | 0.974<br>[0.963, 0.984] | 0.080<br>[0.077, 0.082] | 0.023<br>[0.022, 0.025] | 0.993<br>[0.990, 0.996] | 3.24<br>[2.22, 5.36] |
| Machine features | 0.682<br>[0.664, 0.700] | 0.046<br>[0.041, 0.052] | 0.340<br>[0.309, 0.373] | 0.859<br>[0.856, 0.863] | 0.052<br>[0.046, 0.058] | 0.983<br>[0.981, 0.984] | 3.04<br>[2.64, 3.50] |
| Composite |  |  |  |  |  |  |  |
| WF Model | 0.717<br>[0.707, 0.728] | 0.162<br>[0.152, 0.175] | 0.845<br>[0.830, 0.859] | 0.420<br>[0.415, 0.426] | 0.093<br>[0.089, 0.097] | 0.975<br>[0.972, 0.977] | 3.68<br>[3.317, 4.11] |
| Fusion model | 0.835<br>[0.827, 0.843] | 0.304<br>[0.286, 0.323] | 0.855<br>[0.841, 0.868] | 0.645<br>[0.640, 0.650] | 0.145<br>[0.140, 0.151] | 0.984<br>[0.983, 0.986] | 9.28<br>[8.32, 10.40] |
| RCRI | 0.668<br>[0.657, 0.678] | 0.114<br>[0.107, 0.121] | 0.516<br>[0.496, 0.536] | 0.736<br>[0.731, 0.741] | 0.121<br>[0.115, 0.127] | 0.956<br>[0.953, 0.958] | 2.73<br>[2.52, 2.94] |
| Elixhauser comp. | 0.773<br>[0.763, 0.783] | 0.256<br>[0.239, 0.274] | 0.831<br>[0.816, 0.845] | 0.520<br>[0.515, 0.526] | 0.109<br>[0.105, 0.113] | 0.978<br>[0.975, 0.980] | 4.85<br>[4.38, 5.37] |
| Hannun et al.<sup>33</sup> | 0.712<br>[0.702, 0.723] | 0.150<br>[0.140, 0.161] | 0.809<br>[0.793, 0.825] | 0.474<br>[0.469, 0.479] | 0.098<br>[0.094, 0.102] | 0.972<br>[0.970, 0.975] | 3.54<br>[3.21, 3.92] |
| Ribiero et al.<sup>34</sup> | 0.685<br>[0.674, 0.696] | 0.145<br>[0.135, 0.156] | 0.752<br>[0.735, 0.769] | 0.499<br>[0.493, 0.504] | 0.096<br>[0.092, 0.100] | 0.966<br>[0.963, 0.969] | 2.82<br>[2.59, 3.09] |
| PreOpNet<sup>35</sup> | 0.674<br>[0.663, 0.684] | 0.121<br>[0.114, 0.129] | 0.999<br>[0.998, 1.000] | 0.003<br>[0.002, 0.003] | 0.066<br>[0.064, 0.069] | 0.979<br>[0.948, 1.000] | 3.204<br>[1.25, 5.16] |
| Machine<br>features | 0.651<br>[0.640, 0.663] | 0.116<br>[0.109, 0.125] | 0.744<br>[0.726, 0.761] | 0.461<br>[0.455, 0.466] | 0.089<br>[0.085, 0.093] | 0.962<br>[0.959, 0.965] | 2.34<br>[2.15, 2.56] |

### Cross-validation

Results from 10-fold cross validation are shown in **Table 2** and **Figure 2A-D**. The fusion model demonstrates superior performance across all three outcomes, achieving an AUROC of 0.858 [0.845, 0.872], 0.899 [0.889, 0.908], and 0.835 [0.827, 0.843] for MI, IHM, and the composite outcome, respectively. This performance is significantly better than the WF model (MI: *p* = 1 × 10^−4^; IHM: *p* < 10^−4^; composite: *p* < 10^−4^) and RCRI (MI: *p* < 10^−4^) across all three outcomes. We find the WF model performs well for predicting MI and IHM (MI: 0.791 [0.771, 0.809]; IHM: 0.761 [0.744, 0.777]) and moderately for the composite outcome (0.717 [0.707, 0.728]). While the WF model has a higher absolute performance than RCRI across all three outcomes, this difference is only significant for the composite outcome (MI: *p* = 0.150; IHM: *p* = 0.807; composite: *p* = 1 × 10^−4^).

### Model comparisons

We compare our WF model to three other CNN-based models, from Ribeiro et al.^34^, Hannun et al.^33^, and PreOpNet^35^, trained using our paradigm and data (Table 2). We find that our model performs similarly to Hannun et al.^33^, outperforming slightly over all outcomes, but not significantly (MI: *p* = 0.076; IHM: *p* = 0.891; composite: *p* = 0.897). Our model significantly outperforms Ribeiro et al.^34^ on MI (*p* = 0.013) and composite (*p* = 0.006), but not IHM (*p* = 0.409). It also significantly outperforms PreOpNet^35^ across all outcomes (*p* < 10^−4^). In addition, we compare our model to a non-CNN based model, using 9 ECG features extracted via the Marquette 12SL algorithm^40^ and input into an XGBoost^41^ classifier. We find the WF model outperforms hand-crafted features across all outcomes (*p* < 10^−4^).

### Subgroup analysis

We then take a more granular examination of our model’s performance. Specifically, we begin with a comparison of our models with the current SOC benchmark, the RCRI, in terms of its ability to better differentiate between high- and low-risk patients. We then conduct a subgroups analysis where we stratify patients based on demographics (sex, race, age), admission type (emergent vs. non-emergent), and automated ECG classification based on a commercial software.

### RCRI

We compute the NRI between our models and the RCRI. For MI, we find an NRI of 0.412 [0.234, 0.587] and 0.706 [0.537, 0.867] for the WF and fusion models, respectively. For IHM, we find a lower NRI for both the WF model (0.174 [0.033, 0.316]) and the fusion model (0.614 [0.481, 0.746]). The NRI for the composite model is similar between the WF and fusion models, with values of 0.267 [0.210, 0.324] and 0.292 [0.237, 0.347], respectively. These results indicate that our models outperform the RCRI benchmark in reclassifying patients into appropriate risk categories. A cross-comparison table can be found in **Table S3**.

### Demographics

We compare model performance based on sex, race (White vs. non-White), and age (above or below age 60). We find no significant differences in performance on the basis of sex for the WF (MI: *p* = 0.114; IHM: *p* = 0.080; composite: *p* = 0.053) or fusion (MI: *p* = 0.316; IHM: *p* = 0.461; composite: *p* = 0.058) models. The same is true for race, where we find no significant differences in the WF (MI: *p* = 0.557; IHM: *p* = 0.801; composite: *p* = 0.221) or fusion (MI: *p* = 0.814; IHM: *p* = 0.544; composite: *p* = 0.394) performance between White and non-White subjects. For age, however, we find that the WF model demonstrates significantly higher performance for patients under the age of 60 (MI: 0.831 [0.779, 0.879]; IHM: 0.823 [0.792, 0.853]; composite: 0.718 [0.693, 0.743]) for MI (*p* = 0.005) and IHM (*p* < 10^−4^), but not the composite (*p* = 0.802), as compared to patients over 60 (MI: 0.760 [0.739, 0.781]; IHM: 0.732 [0.713, 0.752]; composite: 0.714 [0.702, 0.727]). The fusion model shows significantly better performance for younger subjects as well (MI: 0.897 [0.864, 0.927]; IHM: 0.920 [0.900, 0.938]; composite: 0.851 [0.834, 0.866]) across all three outcomes (MI: *p* = 0.009; IHM: *p* = 0.047; composite: *p* = 0.007), as compared to older ones (MI: 0.808 [0.789, 0.826], IHM: 0.882 [0.870, 0.893]; composite: 0.817 [0.807, 0.826]). While sex and race do not significantly impact model performance, age appears to be a critical factor, with models performing better for younger patients.

### Admission type

While risk stratification is important regardless of surgery urgency, for non-emergent surgeries, there is significant potential for additional preoperative workup and optimization. For this reason, we divide admission types into two categories: emergency (*n* = 15,578) and non-emergency (*n* = 21,503) and compare our model’s performance. Using the WF model, while we do not find a significant difference in AUROC between emergency and non-emergency surgeries for MI prediction (*p* = 0.963), we do find significantly higher performance for predicting IHM (*p* = 0.010) and composite (*p* = 0.0001) for non-emergent patients. For the fusion model, we achieve significantly higher performance (*p* < 10^−4^) in predicting outcomes for non-emergency surgeries (MI: 0.875 [0.852, 0.896]; IHM: 0.929 [0.912, 0.945]; composite: 0.841 [0.825, 0.857]) as compared to emergency ones (MI: 0.786 [0.763, 0.808]; IHM: 0.835 [0.819, 0.850]; composite: 0.761 [0.750, 0.773]) across all three outcomes. This may indicate the model is particularly valuable in non-emergency or elective surgeries, when risk stratification is most impactful because of the greater latitude for preoperative interventions to reduce complication risk.

### Automated ECG reports

The 12-lead ECG recording devices in this study have an embedded algorithm (Marquette 12SL) which generates automated reports of abnormal findings. We speculated that a computational WF model could extract more specific information than that on commercially available products. To establish this, we examine the odds ratio of our model on ECGs coded as “normal” by the commercial software. In other words, we evaluate the odds of an adverse postoperative event in patients with ECGs coded as normal by the commercial software, but high risk by our WF model. We find a significantly higher odds ratio across all three outcomes (MI: 9.420 [2.016, 19.311]; IHM: 6.977 [5.275, 8.793]; composite: 1.702 [1.500, 1.907]). This suggests that even in ostensibly “normal” ECGs, our model can identify non-obvious abnormalities predictive of future adverse events.

### Temporal stratification

In addition to our cross-validation paradigm, we also evaluate our models on a temporally separate cohort. We divide the patients into a temporally separate development cohort (2008-16; 25,717 patients, 33,854 admissions), which is used to train the model, and test cohort (2017-19; 2,944, 3,227 admissions), which is used to evaluate the generalizability of our method in a chronologically different population (**Figure 2E**). The development and test cohorts have significantly different outcomes likelihoods and comorbidity rates (see **Table 1**). The results of this analysis are summarized in **Table 3**.

**Table 3.**
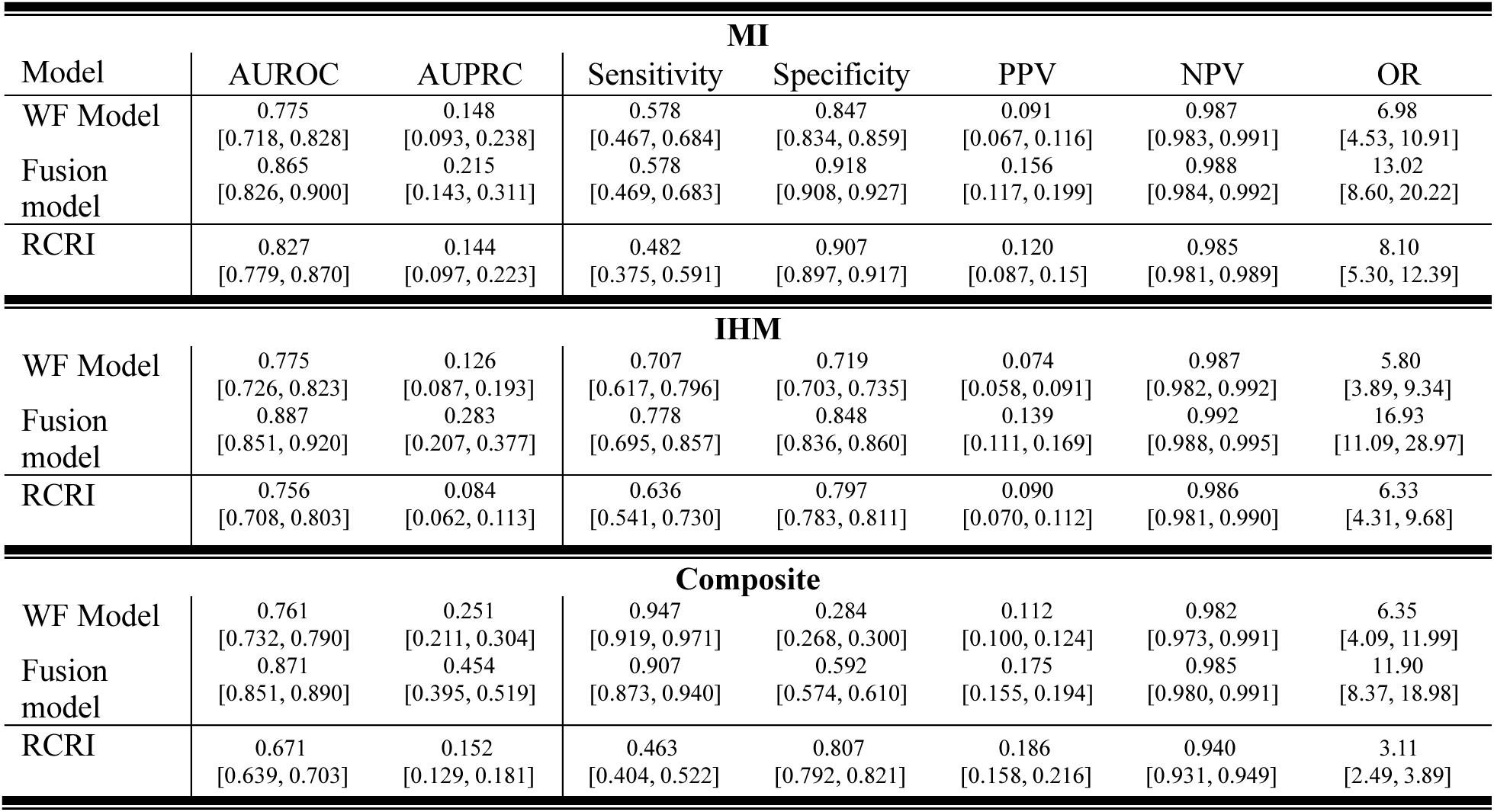
Prospective model comparison. Abbreviated version of the temporal evaluation approach, as in Table 2. Full results can be found in **Table S4**.

When evaluating on the test cohort, our fusion model performs quite well in terms of AUROC (MI: 0.856 [0.826, 0.900]; IHM: 0.887 [0.851, 0.920]; composite: 0.871 [0.851, 0.890]), significantly outperforming the WF model (MI: *p* = 0.049; IHM: *p* = 0.007; composite: *p* < 10^−4^) and RCRI (MI: *p* = 0.002; IHM: *p* = 0.002; composite: *p* < 10^−4^) across all three outcomes. Similar to the cross-validation method, we find that the WF model outperforms RCRI across all outcomes, but this difference is only significant for the composite outcome (MI: *p* = 0.190; IHM: *p* = 0.628; composite: *p* = 0.0002).

### Explainability

One of the challenges in our counterfactual approach to the prediction task is that there is no well-established ground truth for the variations in waveform morphology predictive of adverse postoperative events in noncardiac surgery, as these patients are not actively experiencing severe cardiac complications at the time of prediction (**Figure 3B**). We selected six easily identifiable conditions and provide examples that demonstrate our counterfactual model can learn to reproduce morphological characteristics of each condition (**Figure 3C**).

**Figure 3.**
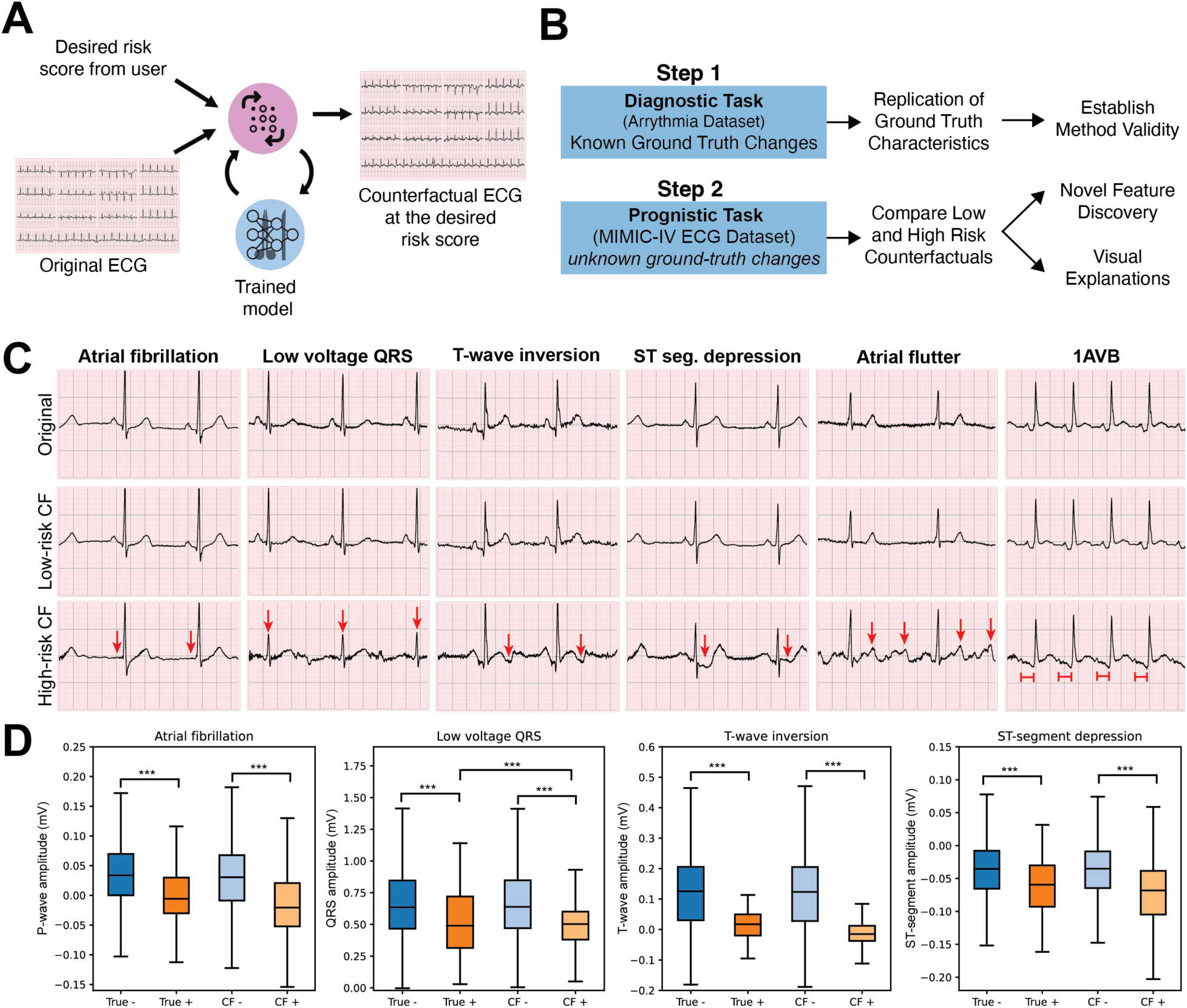
Diagnostic explainability, **(A)** Schematic for the counterfactual generation model. **(B)** Schematic of our counterfactual validation approach. **(C)** Plotted are example ECG strips from the first 2.5 seconds of the waveform from lead II for the conditions labeled above each column of strips. Red arrows indicate morphological changes in the high-risk counterfactuals that are representative of the condition of interest. **(D)** We examine morphological differences relevant to four of the seven conditions of interest (with well-defined and easily extracted morphological features) across the entire diagnostic test set to verify our model reproduces morphological changes characteristic of conditions. “True -” indicates the distribution for patients without the condition, and “True +” indicates the distribution of patients with the condition. Then, for all patients (both true positives and true negatives), we simulate low- and high-risk counterfactuals and extract the corresponding measurements. Significance stars are as follows ∗: *p* < 0.05, ∗∗: *p* < 0.01, ∗∗∗: *p* < 0.001.

In addition to visual examples from single patients, we also conduct a more extensive empirical analysis to validate that our models reproduce the characteristics we expect them to in the diagnostic dataset. To do this, we select four ECG abnormalities (atrial fibrillation, low voltage QRS complex, T-wave inversion, and ST-segment depression) with obvious criteria associated with a single feature that can be automatically extracted, and validate that the presentation is replicated by our counterfactual model. We then delineate each waveform (see **Supplementary Methods**) and extract the average voltage of the T-wave, ST-segment, QRS amplitude, and ST-segment amplitude for each respective condition, as changes in these features are hallmarks of the corresponding conditions. We then compare the values for true positives (e.g., patients with a true diagnosis of T-wave inversion) and true negatives (e.g., patients with no diagnosis of T-wave inversion) and compare these results with our low- and high-risk counterfactuals. We find that our method replicates known changes representative of each of the four common conditions (see **Figure 3D**).

In our exploratory analysis using the *prediction* task of interest, we generate visual examples and conduct a feature correlation analysis. We show how counterfactual ECGs can visually illustrate the morphological differences associated with varying levels of risk for postoperative myocardial infarction. In **Figure 4A**, the low-risk ECG example shows normal QRS duration and standard timing intervals, while the high-risk ECG example exhibits a slightly prolonged QRS duration along with alterations in other morphological features, such as an absence of P waves and moderate depression of the ST-complex. These visual differences highlight the specific ECG changes that our model associates with higher risk, providing interpretable and plausible evidence of the physiological factors contributing to adverse surgical outcomes for a given patient.

**Figure 4.**
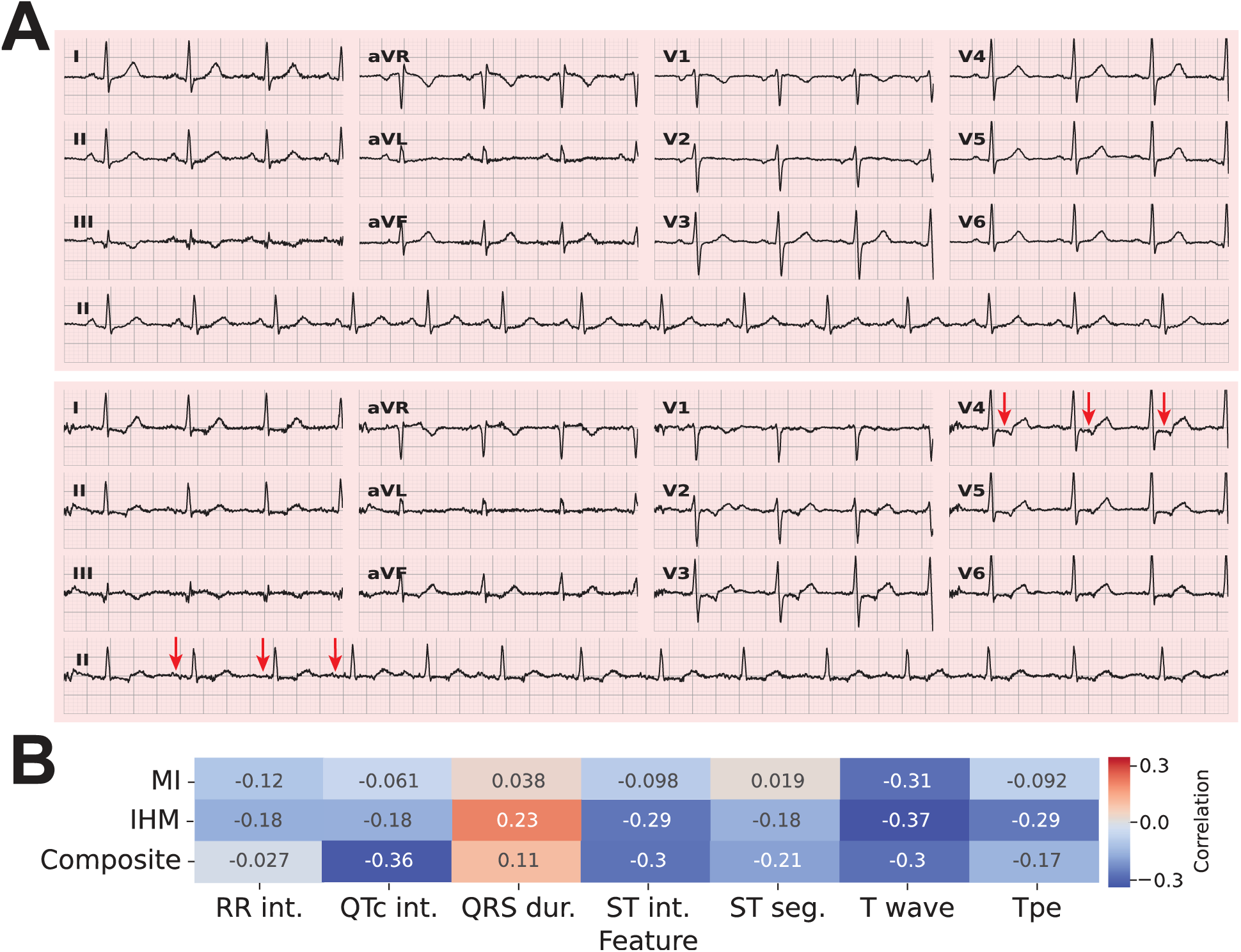
Prognostic explainability. **(A)** Shows low (top) and high (bottom) counterfactual ECG strips for a single patient. Within each strip, the first three subplots show the first 2.5 seconds of electrical activity (with lead numbers inset), and the bottom row shows the full 10-second ECG for lead II. **(B)** Contains a heatmap of the correlation between counterfactual risk score (δ) and each of 7 standard ECG features. See the **Supplementary Information** for a description of each feature.

In our feature analysis, we evaluate the correlation between the counterfactual risk score (i.e., *δ*) and a set of seven ECG timing features (**Figure 4B**). We find a positive correlation between risk score and QRS duration, and a negative correlation between risk score and the other six timing features. This positive correlation between risk score and QRS duration suggests that patients with prolonged QRS complexes are at a higher risk of adverse postoperative outcomes. The QRS duration reflects the time it takes for the ventricles to depolarize and is often prolonged in conditions that indicate cardiac pathology, such as bundle branch blocks or ventricular hypertrophy^43,44^. These conditions can lead to increased myocardial stress or structural abnormalities that predispose patients to adverse events. The negative correlations with the other six timing features may reflect a range of underlying cardiac conditions that accelerate the heart’s electrical activity or alter its recovery phases, potentially increasing the likelihood of complications^45–47^.

## Discussion

We report here on a novel algorithm for preoperative risk stratification developed using a large, publicly available clinical dataset. Our WF model trained on only preoperative 12-lead ECG waveforms outperforms the current benchmark metric (RCRI) across the three outcomes we consider: MI, IHM, and a composite endpoint of stroke, MI, and 30-day mortality. Our WF model significantly outperforms a previously published CNN-based ECG model^35^ for preoperative risk stratification using our training paradigm and data. Next we integrate our waveform-based model with routinely collected clinical variables and demonstrate high performance on our tasks of interest.

The ECG is generally regarded as a diagnostic tool (e.g., for diagnosing pre-existing, or actively occurring conditions). Here, we challenge this view by demonstrating the ECG has significant predictive ability in the perioperative setting. We evaluated our approach on a large cohort of 37,081 hospitalized patients, and on a temporally stratified cohort of 3,227 patients from a chronologically later period. Our model performs well using both paradigms, with our cross-validation fusion model achieving AUROC values 0.835 (composite) and 0.899 (IHM). The fusion model demonstrates similarly high performance in the temporally-stratified group (with AUROCs ranging from 0.865 for MI prediction to 0.887 for IHM), even given significant population differences between the temporally stratified groups (**Table 1**).

We also introduce a novel explanatory approach for waveform prediction models based on the generation of counterfactuals. Most deep learning models are effectively “black boxes,” with complex internal mechanisms that are not readily intelligible. The current standard approach to explainability in deep learning ECG models mostly relies on saliency-based methods, such as locally interpretable model-agnostic explanations^49^ or gradient-based approaches^50^. While these techniques show, roughly, “where” the model is looking, they often fall short of explaining “what” it is looking at – e.g. the specific morphological features influencing the model’s predictions – particularly when these features are subtle or not previously well established. Building on recent generative methods for explainability^51–53^, our research introduces a novel approach using counterfactual explanations to enhance the interpretability of ECG classification models. This method, inspired by foundational work in counterfactual reasoning^36,37^, involves generating “counterfactual” ECGs—modified versions of the original ECG that are minimally altered to change the predicted risk level of an outcome, such as myocardial infarction (MI). By adjusting a control parameter, *δ*, which represents the desired risk level, our model can produce ECGs that not only differ in their risk prediction but also highlight the specific changes in waveform morphology responsible for the different predictions^54^. This approach allows a visual and empirical exploration of how various morphological features contribute to the model’s decisions, offering a more nuanced understanding than what is provided by existing methods.

Notwithstanding, this work has limitations. The lack of external validation raises questions of generalizability across different demographic and clinical settings. However, we note that our model continues to perform well in a temporally distinct sample, despite statistically significant differences in outcome prevalence and comorbidities between development and test cohorts (**Table 1**). The reliance on ICD codes for outcome identification could introduce biases due to inaccuracies in coding practices. As highlighted in various studies^55,56^, the veracity of ICD coding can be compromised by multiple factors (e.g., experience of medical record coders, incomplete physician documentation, etc.), potentially affecting the fidelity of our training labels. However, we note that our model performs similarly for MI and IHM; while the former may be subject to label noise, the latter is not, given the unambiguity of mortality and importance of accurate reporting. This suggests that our models capture latent features within the ECG waveform that are predictive of patient outcomes.

Taken together, these findings indicate that for patients undergoing noncardiac surgery, the 12-lead ECG is a powerfully discriminative prognostic tool. Waveform-based predictive features are not immediately recognizable and can be revealed using deep learning algorithms. Future research should focus on external validation of this model and prospective implementation in real-world clinical settings, which we are currently exploring.

## Data and code availability

We will publicly release the code relevant to replicating the core analyses upon acceptance of the manuscript. The MIMIC-IV data is publicly available.

## Acknowledgements

This material is based upon work supported by the National Science Foundation Graduate Research Fellowship under Grant No. DGE2139757, awarded to CH and SS. Any opinion, findings, and conclusions or recommendations expressed in this material are those of the authors(s) and do not necessarily reflect the views of the National Science Foundation.

## Supplementary Methods

### Baseline methods

#### Elixhauser comorbidity index

The following conditions are used as Elixhauser components, with ICD codes identified by Quan et al.^28^: valvular disease, pulmonary circulation disorders, peripheral vascular disorders, hypertension (with/without complication), paralysis, other neurological disorders, chronic pulmonary disease, diabetes (with and without complication), hypothyroidism, renal failure, liver disease, peptic ulcer disease excluding bleeding, lymphoma, metastatic cancer, solid tumor without metastasis, rheumatoid arthritis/collagen vascular diseases, obesity, weight loss, fluid and electrolyte disorders, deficiency anemia, drug abuse, alcohol abuse, psychoses, depression.

#### RCRI components

Three of the RCRI components (preoperative creatinine, preoperative insulin and a diagnosis of diabetes, and high-risk surgery) are identified using data from the patient’s current hospital stay (i.e., the one for which we are estimating postoperative risk). To identify each of these components, we examine whether the patient has an elevated (> 2 mg/dL) creatinine lab value, a diagnosis of diabetes and a medication order for insulin, or an ICD-9/10 procedure code for intraperitoneal, intrathoracic, suprainguinal vascular surgery, respectively. For the three components based on past medical history (ischemic heart disease, congestive heart failure, or cerebrovascular disease), patients were positively coded for each of the three components if they had any diagnosis in their prior hospital record indicating the presence of the corresponding condition, identified by ICD codes^57^.

#### Machine features

For each ECG in the MIMIC-IV dataset, there is a set of nine real-valued ECG measurements output by the recording software: `rr_interval, p_onset, p_end, qrs_onset, qrs_end, t_end, p_axis, qrs_axis, t_axis`. As a comparison point to our models, we use these features as input to an XGBoost classifier^41^, using the same dataset and evaluation scheme as our non-deep learning models.

### ECG features

To derive our hand-crafted features (e.g., QRS duration, R-wave amplitude, etc.) we used the neurokit2 package^58^. We used lead II to segment ECG peaks (the QRS complex, as well as P- and T-peaks) and their onsets and offsets. We then used the time stamps from lead II to derive our timing variables: RR interval, PR-segment, PR-interval, ST-segment, QT interval, and the durations of the QRS complex, T-wave, and P-wave. Each of these quantities was calculated, and the average duration was used as the feature for that ECG. A description of each feature is included in Table 4.

**Table 4.** Description of hand-crafted ECG features used in counterfactual analysis.

| Feature | Explanation |
| --- | --- |
| RR interval | Time elapsed between R peaks |
| QTc interval | Corrected QT interval. The QT interval is calculated as the time from the start of the Q wave to the end of the T wave. It is corrected to adjust for heart rate:<br>$QT/\sqrt{RR\ interval}$ |
| QRS duration | Time from the start of the Q wave to the end of the S wave. |
| ST interval | Time from the J point to the end of the T wave. |
| ST segment | Duration of the T wave (i.e., start to end). |
| T wave | Duration of the T wave (i.e., start to end). |
| Tpe | Interval from the peak to the end of the T wave. |

### Net reclassification index

We use the net reclassification index^59^ to compare how well our WF and fusion models reclassify subjects, as compared to RCRI. The NRI measures the improvement in risk prediction by assessing the correct reclassification of patients into more accurate risk categories by the new model compared to the baseline model. Specifically, the NRI is calculated based on the proportion of patients with events who are correctly moved to higher risk categories (event NRI) and the proportion of patients without events who are correctly moved to lower risk categories (nonevent NRI). The total NRI combines these components and is defined as the sum of increases in predicted risk among event cases and decreases among nonevent cases. Formally, if we let *u* denote a patient who was “up” classified (i.e., placed into a higher risk category, relative to RCRI), *d* a patient who was “down” classified (i.e., placed into a lower risk category), and *e* the event of interest (e.g., MI), then NRI is calculated as follows^31,60^:

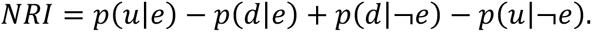

### Hyperparameter optimization

We employ a modified nested k-fold validation scheme to select hyperparameters. We first divide the data into 10 splits, corresponding to the same splits as our main analysis, shown in **Figure 2A**. Then, for each split number *i* ∈ {1, 2, …, 10}, we exclude all the subjects in split i and do not use this in any evaluation. Next, we randomly select 20% of the data to evaluate our hyperapameters on, and using the remaining 80% to train the model. In this way, we preserve the integrity of our k-fold validation approach in the main analysis, by excluding test fold data on each iteration of our nested k-fold validation scheme. We conduct a grid search over 7 hyperparameters (pool size, filter number, dense units, dropout rate, and use of residual/spatial layer), shown in **Table S2**. In total, there are 1080 combinations in our grid search, and we select the hyperparameters with the highest average performance (in terms of AUROC) across all three outcomes.

### Counterfactual generation

To generate counterfactual ECGs, we adopt a method inspired by Explanation by Progressive Exaggeration^36^, which leverages generative adversarial networks (GANs) to generate synthetic ECGs. Our approach is specifically designed for 1D ECG waveforms, as opposed to the 2D image-based approach described in related works^37^. The objective is to produce alternative ECG waveforms that satisfy three primary criteria: (i) realism, meaning the generated ECGs lie on the manifold of training ECGs, (ii) target classification, where the counterfactuals achieve a desired prediction from the classifier, and (iii) similarity, ensuring the counterfactuals are close to the original ECGs. A thorough description of the method and theory is described in DeGrave et al.^37^ and Singla et al.^36^, we provide a brief overview here.

Let *X* ∈ [5000,12] represent an ECG waveform (5,000 time steps, 12 ECG leads) drawn from the data manifold ℳ_*X*_. We define a classifier *f*: *X* → [0,1] that predicts the likelihood of an adverse outcome (i.e., MI, IHM, composite). Our goal is to design a generator *G*: *X* × *C* → [0,1]^*d*^ that outputs a counterfactual waveform 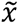 given an input ECG *x* and a condition *c* ∈ *C*, indicating the desired prediction output by the classifier. The conditions c are discrete values indexing bins in the classifier’s output space, defined as *C* = {0,1, … 9} with target outputs corresponding to 10 bins equally spaced on the range from 0 (no risk of adverse outcome) to *r_max_*, where *r_max_* is the maximum risk score to generate (here, 1 for the diagnostic task and 0.2 for the prognostic one; we select a relatively low maximum risk for the prognostic task since our prognostic outcomes are quite rare). The requirements translate to: (i) the range of the generator, *G*(*X*, *C*), must lie within the data manifold ℳ_*X*_, (ii) the classifier’s prediction on the generated waveform, *f*(*G*(*x*, *c*)), should match the target output (bin center corresponding to index *c*), and (iii) if *f*(*x*) is within the bin indexed by *c*, then *G*(*G*(*x*, *c*′), *c*) ≈ *x* for all *c*′ ∈ *C*.

To achieve these properties, we optimize the generator *G* alongside a discriminator network *D*: *X* → ℝ, distinguishing real from generated ECGs. The loss functions for the discriminator *L*_*D*_and the generator *L*_*G*_are as follows^37^:

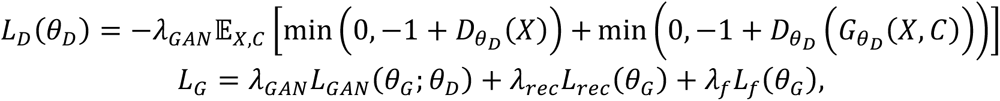

where:

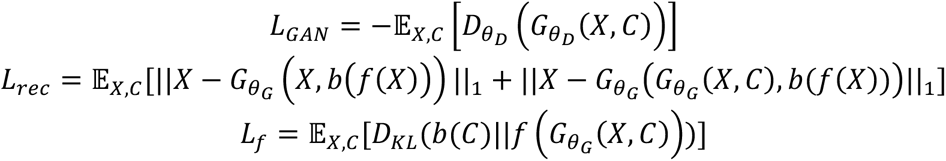

In these equations, *θ*_*D*_ and *θ*_*G*_ are the parameters of the discriminator and generator, respectively. The random variables *X* and *C* are uniformly distributed over *X* and *C*. The function *b*: [0, 1] → *C* returns the bin index of the classifier’s output, and *b*(*C*) ∈ [0, …, *r*_*max*_] returns the center of the bin at index *C*. The Kullback–Leibler divergence is denoted as *D_KL_*. Our generator architecture is based on a residual network-based autoencoder similar to those used in CycleGANs^61^, using code from DeGrave et al.^37^ adapted to 1D convolutional layers for ECG data. We trained our models using an Adam optimizer with a variable learning rate (initial rate of 1 × 10^−4^, which dropped by a factor of 0.1 every 100 epochs) for a total of 400 epochs on a NVIDIA RTX A5500 GPU. We applied spectral normalization^62^ to the discriminator and set *λ*_*GAN*_ = 10, *λ*_*f*_ = 1, and *λ*_*rec*_ = 10.

## Supplementary figures

**Figure S1.**
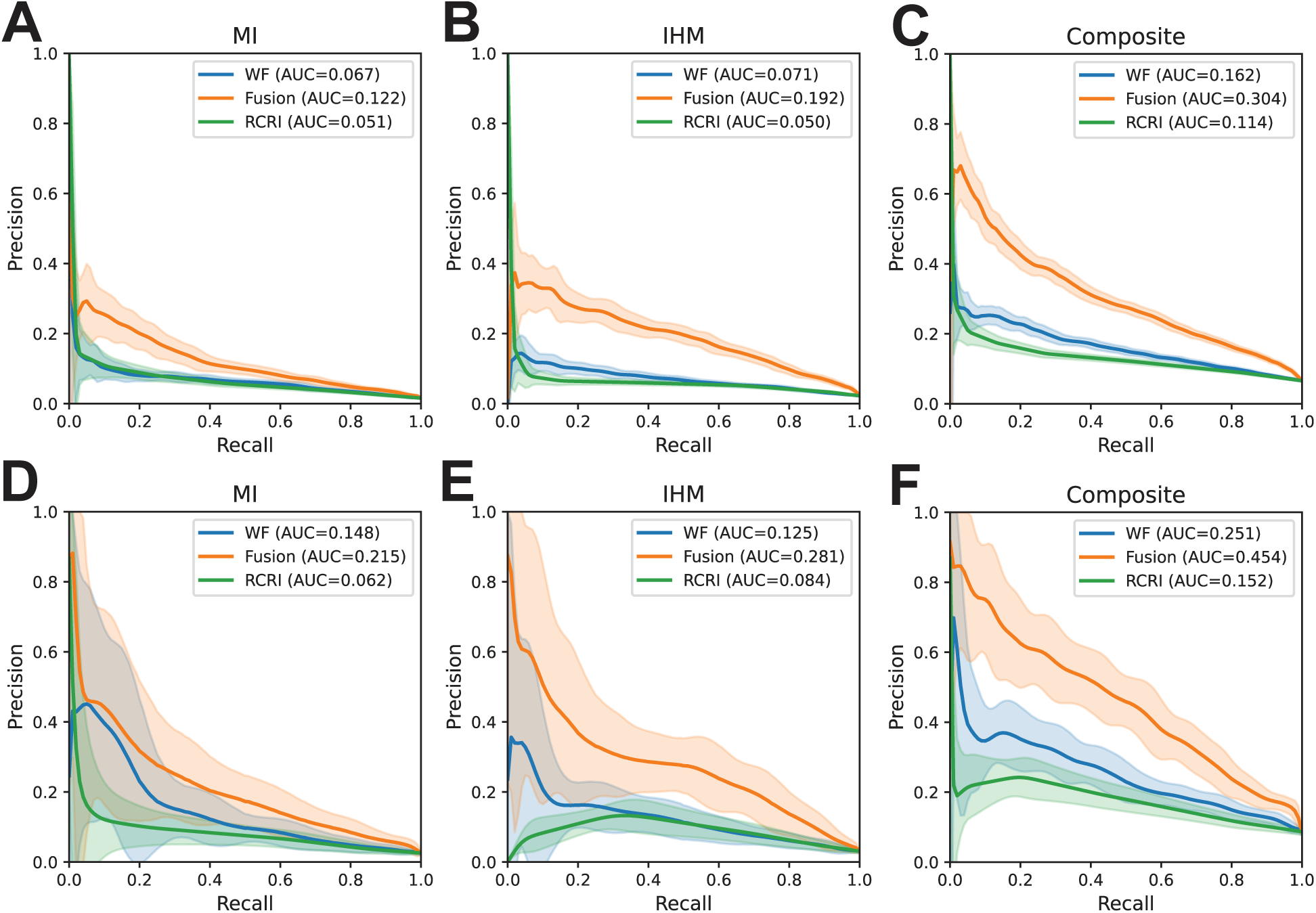
Model PRC. PRC curves for the 10-fold validation scheme for **(A)** MI, **(B)** IHM, and **(C)** composite outcome. **(D-F)** show the corresponding curves for the temporally stratified cohort.

**Figure 6.**
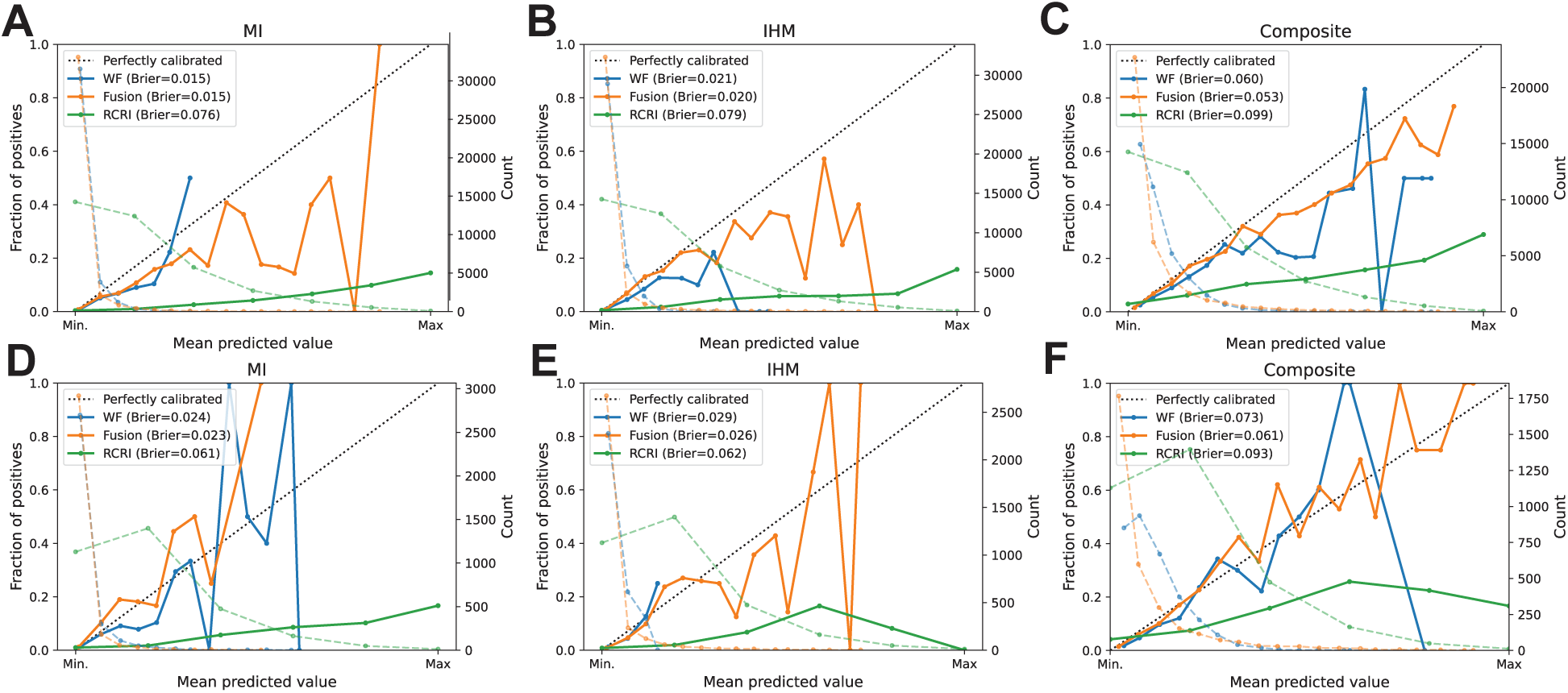
Model calibration. Shown are calibration curves from the 10-fold cross validation **(A-C)** and temporally stratified **(D-F)** cohorts. Inset is the Brier calibration score^63^.

## Supplementary tables

**Table S1.**
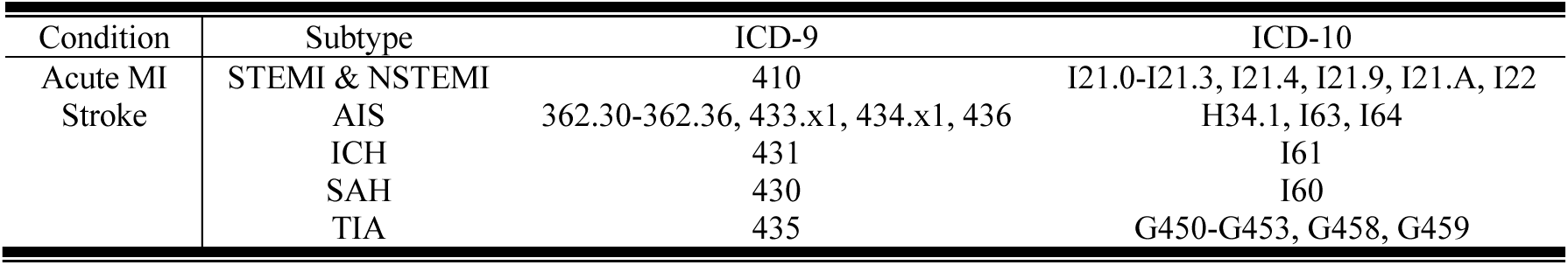
ICD outcome codes. Shown are the ICD-9/10 codes for our two diagnosis-based outcomes (stroke and MI). “Subtype” denotes the subset of the condition the codes in the row correspond to (e.g., a patient has a positive diagnosis of “stroke” if they have one or more of AIS, ICH, SAH, and TIA). Subtype definitions are as follows: STEMI (ST-segment elevation MI), NSTEMI (non ST-segment elevation MI), AIS (arterial ischemic stroke), ICH (intracerebral hemorrhage), SAH (subarachnoid hemorrhage), and TIA (transient cerebral ischemia). Codes for stroke are provided by Kokotailo and Hill^64^. Codes for MI via Clinical Classification Software released by the Healthcare Cost and Utilization Project (HCUP) from the Agency for Healthcare Research and Quality (AHRQ)^23^.

**Table S2.** Grid search parameters.

| Hyperparameter | Values |
| --- | --- |
| Pool size | [2, 2, 4, 2, 2, 4] |
|  | [2, 2, 2, 2, 2, 2] |
| Filter numbers | [8, 8, 16, 16, 32, 32] |
|  | [16, 16, 32, 32, 64, 64] |
|  | [32, 32, 64, 64, 128, 128] |
| Kernel widths | [3, 3, 3, 3, 3, 3] |
|  | [5, 5, 5, 5, 5, 5] |
|  | [7, 7, 7, 7, 7, 7] |
|  | [5, 5, 5, 3, 3, 3] |
|  | [7, 7, 5, 5, 3, 3] |
| Use of residual layer | True, False |
| Use of spatial layer | True, False |
| Dropout rate | 0.1, 0.2, 0.5 |
| Dense units | [64, 32] |
|  | [128, 64] |
|  | [256, 128] |

**Table S3.** RCRI comparison. Shown in each cell is the proportion of adverse outcomes and number of patients (in parentheses), separated by whether the ML and RCRI scores are high and low. The “Avg.” columns and rows denote the column and row average outcome and total patients. For example, the entry 0.081 (3,167) under WF columns high and RCRI high denotes that there were 3,167 patients with an RCRI ≥ 2 and WF risk score ≥ 0.05, of whom 8.1% experienced a postoperative MI.

|  |  | MI |  |  |  |  |  |
| --- | --- | --- | --- | --- | --- | --- | --- |
|  |  | WF |  |  | Fusion |  |  |
|  |  | High | Low | Avg. | High | Low | Avg. |
| RCRI | High | 255 / 3167<br>(0.081) | 157 / 7238<br>(0.022) | 412 / 10405<br>(0.040) | 282 / 3037<br>(0.093) | 130 / 7368<br>(0.018) | 412 / 10405<br>(0.040) |
|  | Low | 71 / 2366<br>(0.030) | 101 / 24310<br>(0.004) | 172 / 26676<br>(0.006) | 57 / 960<br>(0.059) | 115 / 25716<br>(0.004) | 172 / 26676<br>(0.006) |
|  | Avg. | 326 / 5533<br>(0.059) | 258 / 31548<br>(0.008) | 584 / 37081<br>(0.016) | 339 / 3997<br>(0.085) | 245 / 33084<br>(0.007) | 584 / 37081<br>(0.016) |
|  |  | IHM |  |  |  |  |  |
|  |  | High | Low | Avg. | High | Low | Avg. |
| RCRI | High | 331 / 4016<br>(0.082) | 210 / 6389<br>(0.033) | 541 / 10405<br>(0.052) | 450 / 3023<br>(0.149) | 91 / 7382<br>(0.012) | 541 / 10405<br>(0.052) |
|  | Low | 148 / 4160<br>(0.0356) | 134 / 22516<br>(0.006) | 282 / 26676<br>(0.011) | 148 / 1766<br>(0.0084) | 134 / 24910<br>(0.005) | 282 / 26676<br>(0.011) |
|  | Avg. | 479 / 8176<br>(0.059) | 344 / 28905<br>(0.012) | 823 / 37081<br>(0.022) | 598 / 4789<br>(0.125) | 225 / 32292<br>(0.007) | 823 / 37081<br>(0.022) |
|  |  | Composite |  |  |  |  |  |
|  |  | High | Low | Avg. | High | Low | Avg. |
| RCRI | High | 1147 / 8242<br>(0.139) | 114 / 2163<br>(0.053) | 1261 / 10405<br>(0.121) | 1167 / 7025<br>(0.166) | 94 / 3380<br>(0.028) | 1261 / 10405<br>(0.121) |
|  | Low | 919 / 13896<br>(0.066) | 265 / 12780<br>(0.021) | 1184 / 26676<br>(0.044) | 923 / 7368<br>(0.125) | 261 / 19308<br>(0.014) | 1184 / 26676<br>(0.044) |
|  | Avg. | 2066 / 22138<br>(0.093) | 379 / 14943<br>(0.025) | 2445 / 37081<br>(0.066) | 2090 / 14393<br>(0.145) | 355 / 22688<br>(0.016) | 2445 / 37081<br>(0.066) |

**Table S4.** Full prospective model comparison.

| MI |  |  |  |  |  |  |  |
| --- | --- | --- | --- | --- | --- | --- | --- |
| Model | AUROC | AUPRC | Sensitivity | Specificity | PPV | NPV | OR |
| WF Model | 0.775<br>[0.718, 0.828] | 0.148<br>[0.093, 0.238] | 0.578<br>[0.467, 0.684] | 0.847<br>[0.834, 0.859] | 0.091<br>[0.067, 0.116] | 0.987<br>[0.983, 0.991] | 6.98<br>[4.53, 10.91] |
| Fusion model | 0.865<br>[0.826, 0.900] | 0.215<br>[0.143, 0.311] | 0.578<br>[0.469, 0.683] | 0.918<br>[0.908, 0.927] | 0.156<br>[0.117, 0.199] | 0.988<br>[0.984, 0.992] | 13.02<br>[8.60, 20.22] |
| RCRI | 0.827<br>[0.779, 0.870] | 0.144<br>[0.097, 0.223] | 0.482<br>[0.375, 0.591] | 0.907<br>[0.897, 0.917] | 0.120<br>[0.087, 0.157] | 0.985<br>[0.981, 0.989] | 8.10<br>[5.30, 12.39] |
| Elixhauser comp. | 0.827<br>[0.779, 0.870] | 0.144<br>[0.097, 0.223] | 0.482<br>[0.375, 0.591] | 0.907<br>[0.897, 0.917] | 0.120<br>[0.087, 0.157] | 0.985<br>[0.981, 0.989] | 8.10<br>[5.30, 12.39] |
| Hannun et al. <sup>33</sup> | 0.762<br>[0.705, 0.814] | 0.091<br>[0.062, 0.143] | 0.470<br>[0.362, 0.576] | 0.853<br>[0.840, 0.865] | 0.078<br>[0.056, 0.102] | 0.984<br>[0.979, 0.988] | 4.82<br>[3.13, 7.36] |
| Ribiero et al. <sup>34</sup> | 0.739<br>[0.685, 0.790] | 0.079<br>[0.052, 0.132] | 0.494<br>[0.386, 0.602] | 0.834<br>[0.821, 0.847] | 0.073<br>[0.052, 0.095] | 0.984<br>[0.979, 0.989] | 4.61<br>[3.03, 7.08] |
| PreOpNet <sup>35</sup> | 0.658<br>[0.597, 0.716] | 0.049<br>[0.035, 0.078] | 1.000<br>[1.000, 1.000] | 0.002<br>[0.001, 0.004] | 0.026<br>[0.020, 0.031] | 1.000<br>[1.000, 1.000] | Inf<br>[nan, nan] |
| Machine features | 0.585<br>[0.520, 0.651] | 0.047<br>[0.030, 0.090] | 0.241<br>[0.153, 0.337] | 0.882<br>[0.871, 0.893] | 0.051<br>[0.031, 0.075] | 0.978<br>[0.972, 0.983] | 2.30<br>[1.32, 3.67] |
| IHM |  |  |  |  |  |  |  |
| WF Model | 0.775<br>[0.726, 0.823] | 0.126<br>[0.087, 0.193] | 0.707<br>[0.617, 0.796] | 0.719<br>[0.703, 0.735] | 0.074<br>[0.058, 0.091] | 0.987<br>[0.982, 0.992] | 5.80<br>[3.89, 9.34] |
| Fusion model | 0.887<br>[0.851, 0.920] | 0.283<br>[0.207, 0.377] | 0.778<br>[0.695, 0.857] | 0.848<br>[0.836, 0.860] | 0.139<br>[0.111, 0.169] | 0.992<br>[0.988, 0.995] | 16.93<br>[11.09, 28.98] |
| RCRI | 0.756<br>[0.708, 0.803] | 0.084<br>[0.062, 0.113] | 0.636<br>[0.541, 0.730] | 0.797<br>[0.783, 0.811] | 0.090<br>[0.070, 0.112] | 0.986<br>[0.981, 0.990] | 6.33<br>[4.31, 9.68] |
| Elixhauser comp. | 0.842<br>[0.796, 0.883] | 0.179<br>[0.128, 0.251] | 0.667<br>[0.574, 0.758] | 0.867<br>[0.855, 0.879] | 0.137<br>[0.107, 0.168] | 0.988<br>[0.984, 0.992] | 11.37<br>[7.69, 17.55] |
| Hannun et al. <sup>33</sup> | 0.767<br>[0.714, 0.816] | 0.116<br>[0.083, 0.171] | 0.586<br>[0.487, 0.682] | 0.797<br>[0.783, 0.811] | 0.084<br>[0.063, 0.105] | 0.984<br>[0.979, 0.989] | 5.18<br>[3.51, 7.77] |
| Ribiero et al. <sup>34</sup> | 0.724<br>[0.668, 0.774] | 0.113<br>[0.071, 0.174] | 0.505<br>[0.405, 0.602] | 0.759<br>[0.744, 0.774] | 0.062<br>[0.046, 0.079] | 0.980<br>[0.974, 0.985] | 3.07<br>[2.06, 4.52] |
| PreOpNet <sup>35</sup> | 0.674<br>[0.616, 0.730] | 0.071<br>[0.048, 0.106] | 0.909<br>[0.848, 0.962] | 0.152<br>[0.139, 0.164] | 0.033<br>[0.026, 0.040] | 0.981<br>[0.968, 0.992] | 1.76<br>[0.99, 4.38] |
| Machine features | 0.617<br>[0.558, 0.675] | 0.065<br>[0.040, 0.105] | 0.263<br>[0.178, 0.352] | 0.859<br>[0.847, 0.871] | 0.056<br>[0.035, 0.077] | 0.974<br>[0.967, 0.979] | 2.10<br>[1.29, 3.17] |
| Composite |  |  |  |  |  |  |  |
| WF Model | 0.761<br>[0.732, 0.790] | 0.251<br>[0.211, 0.304] | 0.947<br>[0.919, 0.971] | 0.284<br>[0.268, 0.300] | 0.112<br>[0.100, 0.124] | 0.982<br>[0.973, 0.991] | 6.35<br>[4.09, 11.99] |
| Fusion model | 0.871<br>[0.851, 0.890] | 0.454<br>[0.395, 0.519] | 0.907<br>[0.873, 0.940] | 0.592<br>[0.574, 0.610] | 0.175<br>[0.155, 0.194] | 0.985<br>[0.980, 0.991] | 11.90<br>[8.37, 18.98] |
| RCRI | 0.671<br>[0.639, 0.703] | 0.152<br>[0.129, 0.181] | 0.463<br>[0.404, 0.522] | 0.807<br>[0.792, 0.821] | 0.186<br>[0.158, 0.216] | 0.940<br>[0.931, 0.949] | 3.11<br>[2.49, 3.89] |
| Elixhauser comp. | 0.825<br>[0.797, 0.852] | 0.382<br>[0.326, 0.446] | 0.883<br>[0.844, 0.919] | 0.536<br>[0.519, 0.554] | 0.154<br>[0.136, 0.171] | 0.980<br>[0.972, 0.986] | 7.50<br>[5.44, 11.20] |
| Hannun et al. <sup>33</sup> | 0.724<br>[0.695, 0.753] | 0.199<br>[0.167, 0.243] | 0.822<br>[0.776, 0.866] | 0.480<br>[0.462, 0.499] | 0.131<br>[0.116, 0.147] | 0.966<br>[0.956, 0.975] | 3.84<br>[2.90, 5.31] |
| Ribiero et al. <sup>34</sup> | 0.703<br>[0.671, 0.734] | 0.184<br>[0.154, 0.226] | 0.794<br>[0.745, 0.840] | 0.466<br>[0.448, 0.484] | 0.124<br>[0.109, 0.140] | 0.959<br>[0.949, 0.969] | 3.06<br>[2.34, 4.15] |
| PreOpNet <sup>35</sup> | 0.675<br>[0.642, 0.708] | 0.150<br>[0.128, 0.179] | 0.993<br>[0.982, 1.000] | 0.024<br>[0.019, 0.030] | 0.088<br>[0.079, 0.098] | 0.973<br>[0.930, 1.000] | 3.22<br>[1.23, 5.21] |
| Machine features | 0.603<br>[0.567, 0.638] | 0.150<br>[0.121, 0.188] | 0.708<br>[0.654, 0.760] | 0.419<br>[0.401, 0.437] | 0.104<br>[0.090, 0.118] | 0.938<br>[0.924, 0.951] | 1.67<br>[1.31, 2.16] |

